# DIAGNOSTIC ACCURACY OF A NOCTURIA SINGLE QUESTION SCALE AS A PREDICTOR OF SEVERITY OF LOWER URINARY TRACT SYMPTOMS IN MEN

**DOI:** 10.1101/2020.10.05.20206565

**Authors:** Caroline Santos Silva, Carlos Henrique Suzuki Bellucci, Eduardo de Paula Miranda, Mateus Andrade Alvaia, Ricardo Brianezi Tiraboschi, Jose Murillo Bastos Netto, Cristiano Mendes Gomes, José de Bessa Junior

## Abstract

**Objectives:** To evaluated the accuracy of a single question on nocturia to identify men with moderate and severe LUTS.

**Methods:** We evaluated men aged ≥40 years who presented for medical evaluation at two different urological clinics. They completed the IPSS and the Nocturia Single Question Scale (NSQS). NSQS consists of a single question assessing nocturia frequency ranging from 0 to 4. The severity of LUTS using NSQS (index test) versus IPSS (reference standard) was assessed according to the recommendations of the Standards for Reporting of Diagnostic Accuracy Initiative.

**Results:** The accuracy of the NSQS to discriminate patients with severe LUTS based on the ROC curve was 75% (CI 95% 73 – 82%; p<0.001).

**Conclusion:** Patients without nocturia or a single void/night (NSQS <2) have low probability, while NSQS ≥ 3 has a high probability of having moderate or severe LUTS. NSQS is an acceptable alternative to the IPSS, being a fast and simple tool to identify men according to LUTS’s severity.

## INTRODUCTION

Benign prostatic hyperplasia (BPH) is considered one of the most common urological diseases associated with progressive lower urinary tract symptoms (LUTS).^1^ LUTS encompasses a wide range of storage, voiding, and post-micturition symptoms. ^2^. Epidemiological studies have demonstrated that LUTS’s prevalence is over 60% in men aged > 40 years. ^3,4^

Guidelines for BPH management recommend that the International Prostate Symptom Score (IPSS) be routinely used as a standard procure during patient evaluation. (5,6) The IPSS is an 8-item questionnaire that consists of seven symptom questions and one quality of life question. ^7^ It has been validated in many countries and enables the stratification of patients according to the severity of symptoms. This categorization is a useful instrument to determine disease severity, response to therapy, progression, and to guide the management of LUTS in men. ^5,6^

Although the IPSS is helpful in clinical practice, it is lengthy and time-consuming. Simplifying the instruments designed to evaluate LUTS might reduce the burden placed on the respondent, broaden the instrument application, and improve compliance. ^8^ In developing countries, the adoption of IPSS has challenges, such as a few specialist availabilities and little patient contact time. ^9^ Also, emerging data have shown that men with limited literacy and numeracy frequently misunderstand IPSS. ^10^ In this scenario, simplified psychometric instruments to assess urinary symptoms have emerged ^8,11^, but validation is still pending in low literacy and diverse populations.

Recently, it has been demonstrated that a single-question assessment of nocturia might be a useful alternative to the IPSS. ^12^ Single-question questionnaires provide a brief and easily administrated tool for the detection of diseases and have been recommended as a screening tool for different specific medical conditions. ^13–17^ Therefore, we sought to investigate if a nocturia single-question scale is an accurate diagnostic method in the categorization of LUTS severity in a large Brazilian population.

## METHODS

### Study population

We performed a cross-sectional study between July and December 2019. Study participants consisted of men aged ≥40 years who attended regular follow-up appointments in two urologic clinics located in different cities in Brazil. Subjects with active or history of urinary tract infection or within the past month were excluded.

This study was approved by the Research Ethics Committee of our institution, and all participants provided written informed consent.

All methods, definitions, and units are according to the standards recommended by The International Continence Society. ^2^

### LUTS assessment

“Nocturia” was defined as any necessity to void during the main sleeping period, in which each micturition must be preceded and followed by any course or intention to sleep. ^2^.

LUTS were assessed using self-administered IPSS validated in Portuguese. For patients with low literacy levels presenting comprehension difficulties, the questionnaire was administered by a research assistant, who would clear last doubts in meaning or context. The IPSS is composed of 7 questions on urinary symptoms, including the sensation of incomplete emptying, urinary frequency, intermittency, urgency, weak stream, straining, and nocturia. Responses to all but the last question are scored as the following options: none (score 0), < 1 in 5 times (score 1), < half the time (score 2), about half the time (score 3), > half the time (score 4), or almost always (score 5). The last question evaluates the nocturia frequency and is scored in 6 ordered categories from none to 5 or more times. These questions form a scale by summing the responses (0 to 5 for each response), and patients may be categorized as asymptomatic (0 points), mild symptoms (1-7 points), moderate symptoms (8-19 points), and severe symptoms (20-35 points). The NSQS was obtained from the last question of IPSS that evaluates nocturia. It is scored in five ordered categories from none to 4 or more times and was administered independently to all patients.

### Sample size calculation and diagnostic accuracy evaluation

Considering a minimal disease prevalence of 15% according to previous studies evaluating LUTS in the male population, a sample of at least 615 subjects would be necessary for a sensitivity/specificity of 80%, absolute precision of 5%.

We investigated the diagnostic properties of the NSQS (index test) for the categorization of LUTS severity based on the IPSS score (reference standard). The time of completion (in minutes) of each questionnaire was measured to estimate the respondent burden. As a study of accuracy, this article complies with the recommendation of the Standards for Reporting of Diagnostic Accuracy Initiative. ^18^

### Statistical analysis

Data were presented as absolute values, frequencies, or medians and interquartile ranges [IQR]. The sensitivity, specificity, predictive values, and likelihood ratio of each NSQS, including 95% confidence intervals (CI), describes the diagnostic accuracy. Receiver operator characteristic (ROC) curves were produced to visualize and calculate the area under the curve (AUC) that was used to describe the diagnostic characteristics of the NSQS to diagnose the severity of LUTS. Statistical analyses were performed using GraphPad Prism version 8.4.0.

## RESULTS

### Study population

A total of 763 patients were initially enrolled. Thirty-seven patients (4.8%) were excluded from the analysis because of the current or recent history of urinary tract infection, and 29 (3.8%) refused to participate. Thus, 697 (91.4%) men were included in the analyses. Median age was 60.0 [54.0-68.0] years. Median IPSS and of NSQS episodes were 9.0 [5-17] and 2.0 [1-3], respectively.

### Prevalence of LUTS

According to the IPSS, patients presented mild, moderate, and severe symptoms in 262 (37.6%), 279 (40.0%), and 129 (18.6%), respectively. Twenty-seven (3.8%) were asymptomatic.

The NSQS was zero in 118 (17%), one in 141 (20%), two in 197(28%), three in 144(21%), and four or more in 95 subjects (13%). The NSQS was higher in subjects with moderate/severe LUTS than in asymptomatic and with mild symptoms, respectively, 2[2-3] and 1 [0-2]. (p<0.001) (Figure 1)

**FIGURE 1:**
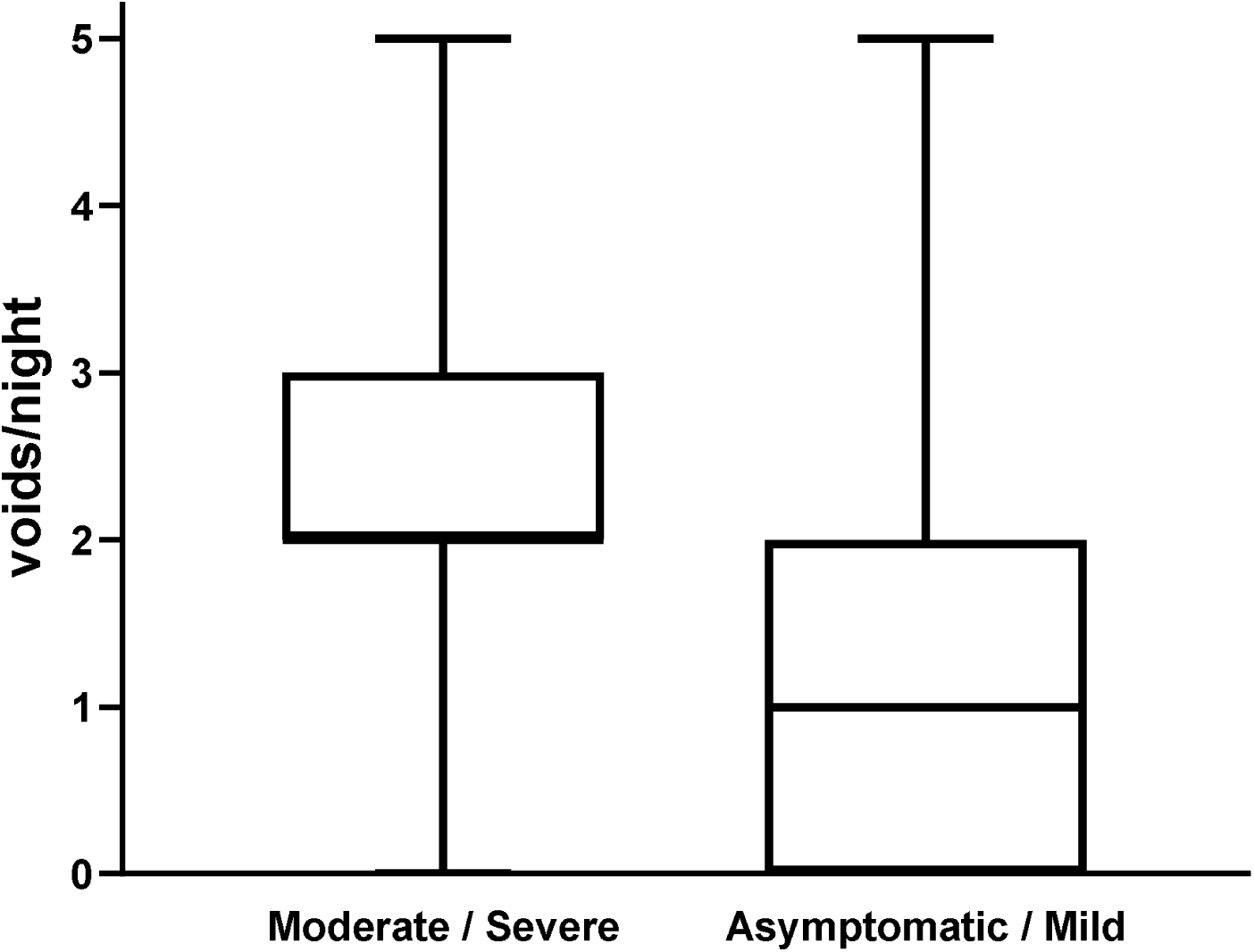
Box-plot describes the number of nocturia episodes in subjects with moderate/severe and in asymptomatic/mild LUTS. LUTS = Lower urinary tract symptoms.

### NSQS diagnostic properties

The overall accuracy of NSQS in discriminate more severe LUTS, estimated by the ROC curve, was 0.75 (95% CI 0.72-0.79) (Figure 1). The sensitivity, specificity, predictive values, the positive and negative likelihood ratio of NSQS of each value are depicted in Table 1.

**TABLE 1:**
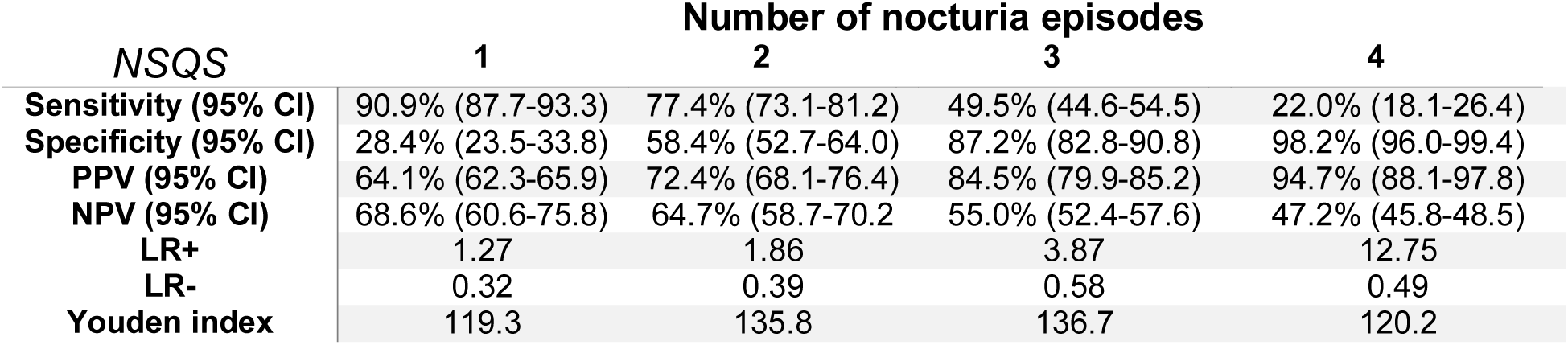
Diagnostic parameters of NSQS thresholds to discriminate patients with mild (IPSS < 8) or moderate/severe (IPSS ≥ 8) LUTS. NSQS = Nocturia single question scale. IPSS = international prostate symptom score, IC = Confidence intervals, PPV = Positive predictive value, NPV = Negative Predictive Value, LR+ = Positive likelihood ratio, LR- = Negative Likelihood Ratio

| NSQS | Number of nocturia episodes |  |  |  |
| --- | --- | --- | --- | --- |
|  | 1 | 2 | 3 | 4 |
| **Sensitivity (95% CI)** | 90.9% (87.7-93.3) | 77.4% (73.1-81.2) | 49.5% (44.6-54.5) | 22.0% (18.1-26.4) |
| **Specificity (95% CI)** | 28.4% (23.5-33.8) | 58.4% (52.7-64.0) | 87.2% (82.8-90.8) | 98.2% (96.0-99.4) |
| **PPV (95% CI)** | 64.1% (62.3-65.9) | 72.4% (68.1-76.4) | 84.5% (79.9-85.2) | 94.7% (88.1-97.8) |
| **NPV (95% CI)** | 68.6% (60.6-75.8) | 64.7% (58.7-70.2) | 55.0% (52.4-57.6) | 47.2% (45.8-48.5) |
| **LR+** | 1.27 | 1.86 | 3.87 | 12.75 |
| **LR-** | 0.32 | 0.39 | 0.58 | 0.49 |
| **Youden index** | 119.3 | 135.8 | 136.7 | 120.2 |

The median time of completion was significantly lower for NSQS than to IPSS, respectively, 0.28 [0.12-0.45] and 2.7 [2.3-3.5] minutes. (p < 0.001).

## DISCUSSION

Lower urinary tract symptoms are prevalent in adult men ^3^, and assessing LUTS is important to the clinical decision process. ^6^ The IPSS is the most used tool to evaluate LUTS severity; however, it’s lengthy, bothersome, and time-consuming. Furthermore, in developing countries, few urologists’ availability, little patient contact time, and low literacy/numeracy have limited the IPSS widespread use. The present study represents an effort to develop a simplified single question instrument to predict LUTS severity in adult men. A study with 407 patients from a county hospital and a university hospital compared answers from interviewer-administrated and self-administrated IPSS and found that only 72% of individuals understood all seven questions. ^19^ In this study, education level had a significant impact on questionnaire understanding, in which patients with lower educational levels were more prone to misunderstand the IPSS, misrepresent their symptoms, and, ultimately, receive inappropriate treatment.^19^ Another study evaluating 70 men from Nigeria demonstrated that only a quarter could understand all the questions of IPSS.^20^ Therefore, in developing countries, shorter and simplified tools may be particularly useful to assess LUTS in adult men, with possible implications on public health.

In the present study, we have demonstrated an interesting accuracy of NSQS in the detection of more severe cases of LUTS. The NSQS presents acceptable accuracy properties to screen the severity of LUTS in men in the primary care setting. Our data demonstrated that 85% of men with NSQS ≥ 3 presents moderate or severe LUTS and that 2/3 of men with NSQS = 0-1 presents IPSS < 8. Two cut-offs (2 and 3 episodes of nocturia per night) are the best and have a very similar overall accuracy, despite significant differences in sensitivity and specificity.

The International Continence Society has recommended that nocturia should be quantified through a bladder diary.^2^ However, previous studies have revealed that nocturia is a LUTS that patients easily recall ^21^, and could be reliably assessed through direct questioning. Therefore, we have evaluated nocturia through a single question scale as our main objective was to test a simple, quick, and reproducible tool. Although well-defined recommendations are supporting the use of bladder diaries in research and clinical practice, it tends to be excessively demanding to the patient population and is considered not in alignment with the purposes of our study.

NSQS ≥ 2 is particularly relevant as it is the standard number of voids considered in the definition of nocturia by most authors. Some studies have consistently demonstrated that this frequency is clinically relevant, bothersome, and associated with reduced quality of life. ^22–24^ Despite its lower specificity, it has good sensitivity and can be used when a more comprehensive evaluation of nocturia is required.

On the other hand, NSQS ≥ 3 has a slightly higher overall accuracy than two episodes by night. It has superior specificity, which provides a high probability of moderate or severe LUTS for those beyond the cut-off, despite having a lower sensitivity. We believe that a higher predictive value could be particularly useful in the primary care setting in our country, which is often characterized by restricted resources and the insufficiency of urologists in the healthcare system.

Although the IPSS is the most recommended tool for the assessment of LUTS severity in BPH patients ^5,6^, its use is potentially limited by its length, which could be a burden to patients that would have to read, understand and answer all the questions. Efforts have been applied to create simplified methods capable of evaluating LUTS in adult men, such as the UWIN (Urgency, Weak stream, Incomplete emptying, and Nocturia) questionnaire. ^25^

Recently, our research group validated the Portuguese version of UWIN in a Brazilian cohort.^26^ UWIN provides comparable results to the IPSS while using a more straightforward format and taking less time to complete. However, it comprises a four questions questionnaire with scores ranging from 0-3, adding up to a composite score of 0-12. The UWIN, although simpler than IPSS, might still add considerable effort from respondents and physicians.

Other abbreviated Patient-Reported Outcome Assessments (PROs) have been proposed to decrease the burden of IPSS on respondents and clinicians, such as QPT (Quick Prostate Test) and FLOW (Frequency, Leakage, Overnight voiding, and Weak stream). ^27,28^ These instruments demonstrated useful equivalence to the IPSS and some advantages, including efficiency and ease of application. In this scenario, we have decided to investigate if a single-question questionnaire could be advantageous over the already abbreviated UWIN.

Single questions have also been recommended for screening of other medical conditions such as depression, dementia, schizophrenia, sedentary behavior, and erectile dysfunction. ^13–17^ Single questions offer a brief, easily administrated screening tool for the detection of diseases. The present study demonstrated that NSQS might be used as a screening tool to predict male LUTS severity (Figure 2). The overall accuracy of NSQS to discriminate between absent/mild or moderate/severe LUTS was 75%.

**FIGURE 2:**
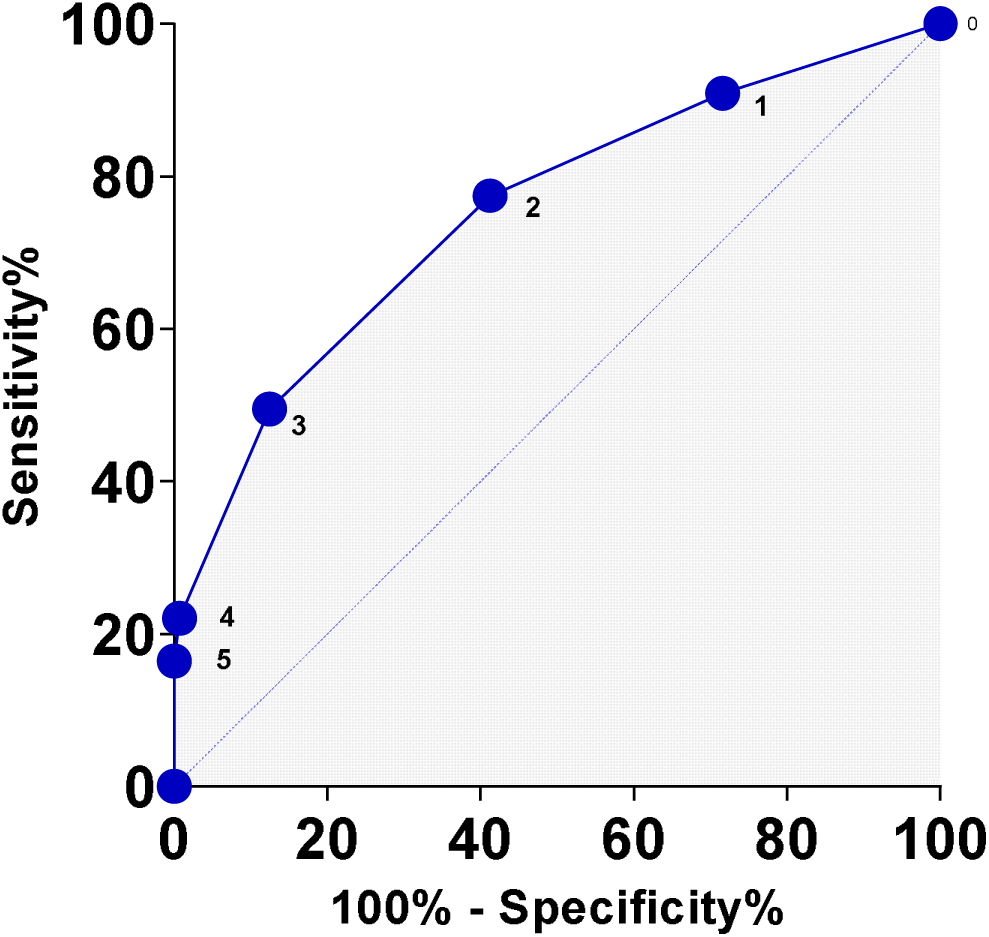
ROC curves of NSQS in distinguishing patients with absent and mild (IPSS < 8) versus moderate and severe (IPSS ≥ 8) LUTS. ROC curves = *Receiver Operating Characteristic Curve*. NSQS = Nocturia single question scale. IPSS = international prostate symptom score. LUTS = Lower urinary tract symptoms.

A previous study has also investigated the role of a single nocturia question in the evaluation of 162 African men ≥ 50 years, comparing the accuracy of NSQS and IPSS.^12^ This study has demonstrated that NSQS ≥ 3 is the most accurate cut-off to indicate bothersome LUTS in men. The sensitivity and specificity of NSQS to discriminate men with mild (IPSS <8) from men with moderate/severe LUTS (IPSS≥8) were 87.0% and 91.0%, respectively. Also, both positive and negative predictive values were 89.0%. In our series, the sensitivity observed was significantly lower in comparison to this African study. Such a difference might be a reflection of sample variations, casuistic characteristics, and potential response bias arising from cultural differences and literacy. Our sample was predominantly urban and apparently with a higher level of education.

The current study has demonstrated that simplified PROS also are accurate tools to predict LUTS severity in adult men without being a time-consuming process to responders and researchers. Future studies should compare the accuracy of these different simplified instruments in different populations, and the usefulness of all these tools to evaluate LUTS severity, response to therapies, and symptom progression.

Interview-based completion of validated questionnaires has also been an alternative to overcome a patient’s limited linguistic comprehension. However, having a medical professional solely to manage questionnaires is certainly not cost-effective. Furthermore, having the attending physician explain each question in detail for this purpose would eventually mean additional workload for the already scarce specialists available in remote and underserved areas.

Although our study presents relevant data to clinical practice, our findings must be interpreted with caution due to some limitations. This study was a nonrandomized cohort of Portuguese-speaking Brazilian men in two tertiary urological centers in an urban region. Furthermore, our population included highly literate individuals, as almost 95% could self-administer the IPSS, which limits validation in other scenarios. Future studies should build on our findings in more diverse clinical settings such as veterans’ hospitals, teaching hospitals, and in the primary care environment.

## CONCLUSIONS

The NSQS may be used as a screening tool to predict the severity of LUTS in men. Patients with none or one void/night have a low probability of severe LUTS, while those with three or more voids have a high probability of moderate or severe symptoms. NSQS can be used as a quick and less expensive alternative in daily clinical practice that could be implemented in the primary care setting.

## Data Availability

The data are in the custody of the senior researcher. They will be available upon request

